# Leveraging Genetic Correlations to Prioritize Drug Groups for Repurposing in Type 2 Diabetes

**DOI:** 10.1101/2025.06.13.25329590

**Authors:** Astrid Johannesson Hjelholt, Tahereh Gholipourshahraki, Zhonghao Bai, Merina Shrestha, Mads Kjølby, Peter Sørensen, Palle Duun Rohde

**Affiliations:** Centre for Quantitative Genetics and Genomics, Aarhus University, Denmark; Steno Diabetes Centre Aarhus, Aarhus University Hospital, Denmark; Department of Clinical Pharmacology, Aarhus University Hospital, Denmark; Department of Endocrinology and Internal Medicine, Aarhus University Hospital, Denmark; The National Centre for Register-based Research, Aarhus University, Denmark; Department of Biomedicine, Aarhus University, Denmark; Genomic Medicine, Department of Health Science and Technology, Aalborg University, Denmark; Department of Clinical Genetics, Aalborg University Hospital, Aalborg, Denmark

## Abstract

Type 2 diabetes (T2D) is a complex, polygenic disease with substantial health impact. Despite extensive genome-wide association studies (GWAS) identifying risk loci, therapeutic translation remains limited. We applied a Bayesian Linear Regression (BLR) multi-trait gene set model to prioritize druggable gene sets, integrating GWAS summary statistics with drug-gene interaction data from the Drug Gene Interaction Database (DGIdb). For each drug group, defined at the ATC 4th level, we calculated posterior inclusion probabilities (PIP) to assess relevance. Known antidiabetic agents showed strong associations with T2D, validating the model. Additionally, carboxamide derivatives, fibrates, uric acid inhibitors, and various immunomodulatory and antineoplastic agents demonstrated significant genetic relevance. Gene-level analyses highlighted key T2D-associated genes, including *PPARG*, *KCNQ1*, *TNF*, and *GCK*. Notably, bezafibrate, a *PPAR* pan-agonist, demonstrated substantial genetic overlap with T2D loci, supporting its potential in metabolic disease. This study introduces a genetically informed pipeline for drug repurposing based on multi-trait gene set analysis.

## Introduction

Type 2 diabetes (T2D) affects over 500 million people globally and continues to rise, posing a major global health challenge [1]. T2D is characterized by insulin resistance and compromised insulin secretion [2], leading to chronic hyperglycaemia and complications such as cardiovascular disease, chronic kidney disease, retinopathy, and neuropathy [3, 4]. Despite important advances in T2D treatment, the development of novel therapies remains a key priority to improve glycemic control, reduce complications, and enable more personalized treatment approaches [5].

The pathogenesis of T2D is multifactorial, involving a combination of lifestyle and environmental factors along with an underlying polygenic predisposition [2, 6]. The heritability of T2D is estimated to be between 40% and 70% [7, 8]. Genome-wide association studies (GWAS), which systematically assess allele frequency differences of common DNA variants between healthy individuals and individuals with the disease of interest [9, 10], have identified more than 600 independent genetic loci associated with T2D risk [11, 12]. Although GWAS have been instrumental in identifying predisposing genetic loci, these individual genomic regions do not fully capture the collective influence of functionally related genes. Gene set enrichment analyses have emerged as a valuable tool for assessing the joint effect of multiple genetic variants within predefined gene sets [13, 14]. In a recent study, we developed a gene set prioritization method using a Bayesian Linear Regression (BLR) model to identify sets of genes associated with a complex trait [15]. Furthermore, by using multi-trait analyses, we were able to analyse related traits jointly, thereby increasing detection power.

Genetically supported drug targets are more likely to succeed in clinical trials, highlighting the potential of GWAS to guide drug discovery [16, 17]. While de novo drug discovery remains an expensive and time-consuming process, drug repurposing offers an efficient alternative by leveraging existing pharmacological and safety data [10]. We propose that statistically genetic informed approaches, such as the BLR multi-trait gene set prioritization model, may enhance drug repurposing efforts by pinpointing genetically validated targets.

The aim of this study, was to identify drug candidates, which may have the potential to be repurposed for the treatment of T2D using the BLR multi-trait model in a genetically informed, drug-repurposing pipeline by combining summary statistics from large GWAS of T2D and related traits with drug-gene interaction sets from the Drug Gene Interaction Database (DGIdb) [18].

## Methods

A genetically informed drug-target prioritization was performed by utilizing a gene set prioritization approach based on a BLR model described and evaluated by Gholipourshahraki et al. [19]. In short, gene-level summary statistics (𝑍-scores) were computed based on GWAS summary statistics for T2D and related traits while accounting for linkage disequilibrium (LD) using the VEGAS (Versatile Gene-Based Association Study) algorithm [20]. A design matrix linking genes to gene sets was constructed to integrate curated gene sets. Gene sets, defined as sets of genes linked to a drug or chemical subgroup (4^th^ level ATC group), were derived from DGIdb [18]. The BLR model was then fitted using this design matrix of all gene sets as input features (predictors) with the gene-level 𝑍-scores as the response variable. For each gene set linked to a drug or ATC group, the posterior inclusion probability (PIP) was obtained from the BLR model to assess the probability of inclusion in the model and the strength of the association. Gene sets with higher PIP-values, indicating a stronger association with T2D, suggest that drugs linked to these gene sets may be useful for treating or managing T2D, thereby facilitating the identification of potential drug targets. By extending the methodology to a multi-trait analysis, a comprehensive exploration of gene sets across diverse traits was enabled. A schematic overview of the workflow is illustrated in Figure 1. Details on the statistical model and analyses and the data used are provided in the following sections.

**Figure 1.**
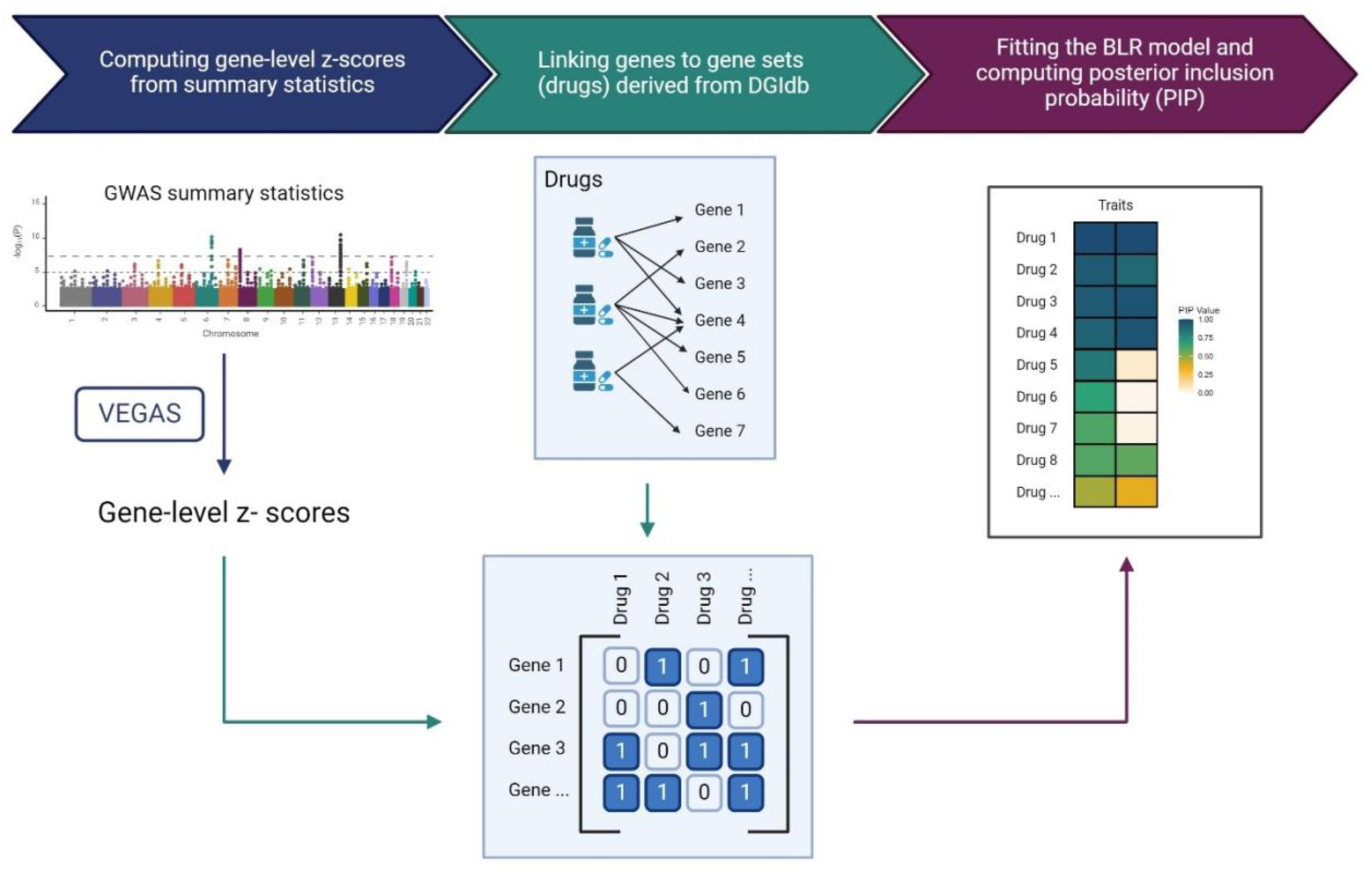
Overview of workflow. Gene-level Z-scores for T2D and related traits were computed using GWAS summary statistics and the VEGAS algorithm. A design matrix linking genes to gene sets was constructed to integrate curated gene sets (drugs linked to sets of genes) derived from DGIdb. The BLR model was then fitted using this design matrix of all gene sets as input features (predictors) and the Z-scores as the response variable. For each gene set (drug), the PIP was computed to determine likelihood of inclusion in the model. VEGAS (Versatile Gene-Based Association Study), DGIdb (Drug Gene Interaction database), posterior inclusion probability (PIP).

### Data processing and integration

Data processing and integration were facilitated by using the R package gact, which is designed to establish and populate a comprehensive database focused on genomic associations with complex traits [21].

### GWAS summary data

We applied the BLR models to nine distinct complex trait phenotypes, all related to T2D, with publicly available GWAS summary data. These include T2D [22], coronary artery disease (CAD) [23], chronic kidney disease (CKD) [24], hypertension (HTN) [25], body mass index (BMI) [26], waist-hip ratio (WHR) [26], glycated haemoglobin (Hb1Ac) [24], height [27], systolic blood pressure (SBP) [28], and triglycerides (TG) [29].

### LD reference data and gene annotation

Reference genotype data from the 1000 Genomes Project [30] (European ancestry) were used to estimate pairwise LD among common variants. Genetic variants with a minor allele frequency below 0.01, a call rate lower than 0.95, and those not conforming to Hardy-Weinberg equilibrium (with a *P*-value of 1 × 10^−12^) were excluded. Additionally, genetic variants exhibiting ambiguous alleles (such as GC or AT), having multiple alleles, or representing indels, were removed [31].

Gene-level markers included variants 35 kb upstream to 10 kb downstream of the coding region, based on Ensemble GRCh37.87 annotations: ftp.ensembl.org/pub/grch37/current/gtf/homo_sapiens/Homo_sapiens.GRCh37.87.gtf.gz.

### Gene sets

Gene sets were derived from multiple annotation sources. Drug-gene interaction sets were obtained from DGIdb [32], while gene–disease associations were retrieved from JensenLab [33], covering text-mined, expert-curated, experimental, and integrated datasets (full and filtered versions of *human_disease_textmining*, *human_disease_knowledge*, *human_disease_experiments*, and *human_disease_integrated* .tsv files). These comprehensive datasets were used to enrich the analysis with evidence-based links to human diseases.

### Statistical models and analyses

Our method is built upon a linear model that utilizes the matrix notation shown below:

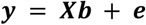

where 𝒚 denotes the per-gene statistic, which measures the association between individual genes and the trait phenotype, 𝑿 is a design matrix linking genes to gene sets and the corresponding per-gene statistic, and 𝒆 represents the residuals, which are presumed to follow an independent and identically distributed normal distribution with a mean of 0 and a variance of 𝜎^2^. The dimensions of 𝒚, 𝑿, 𝒃 and 𝒆 depend on the number of traits (𝑘), gene sets (𝑚), and genes (𝑛). The elements in the design matrix 𝑿 has the value one if a gene is part of a gene set, and zero otherwise. The vector 𝒃 denotes the regression coefficient for each gene set.

### Single trait BLR model

A BLR model was implemented based on the BayesC [34] prior assumptions to model the association between gene sets and traits. The BayesC approach employs a spike-and-slab prior distribution:

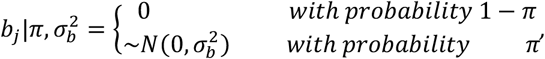

assuming the regression effects (𝒃) follow a mixture distribution, comprising a point mass at zero and a normal distribution defined by a common variance 𝜎_b_^2^ for the regression effects. Each regression effect (𝑏_𝑗_) can either be zero, indicating no effect, or nonzero, indicating its contribution to the response variable. The prior probability, 𝜋 = 0.001 specifies the fraction of regression effects that are expected to belong to each category. The prior distribution of the common variance 𝜎_b_^2^ for the regression effects follows an inverse Chi-square distribution, 𝜒^−1^(𝑆_𝑏_, 𝜈_𝑏_) [35], where 𝑆_𝑏_ represents the scale parameter of an inverse Chi-square distribution and 𝜈_𝑏_ represents the degrees of freedom parameter.

The mixture proportions are determined using a Dirichlet distribution (𝐶, 𝑐 + 𝛼), where 𝐶 represents the number of mixture components in the distribution of regression effects, 𝑐 represents the vector of counts of regression variables within each component, and 𝛼 = (1,1) is the concentration hyperparameters ensuring that the sampled mixture proportions is entirely determined by the information in the data. To simplify computations and analyse these complex distributions, a data augmentation technique is employed. A latent variable, 𝑑 = (𝑑_1_, 𝑑_2_ …, 𝑑_𝑚−1_, 𝑑_𝑚_),), is introduced to indicate whether the 𝑗^𝑡ℎ^ regression effect is zero or nonzero.

### Multi-trait BLR models

We implemented a multi-trait BLR model based on the BayesC prior [34], allowing each gene set to influence any combination of traits. This approach improves detection of shared biological functions across correlated traits by leveraging shared information and applying regularization, as in the single-trait model. For the case of analysing two traits, the core equation for estimating regression effects is given by:

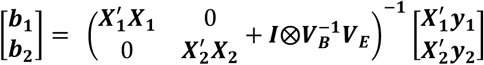

Key parameters in this model include 𝑽_𝑩_, the covariance matrix of the regression effects, and the residual covariance matrix, denoted as 𝑽_𝑬_. These matrices capture the shared relationships between regression effects across traits.

For the two-trait case the covariance matrix 𝑽_𝑩_ is represented as:

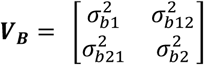

When 𝑽_𝑩_ is not uniform across gene sets, it enables differential shrinkage of gene set effects, implemented through “spike-and-slab” priors. Additionally, if the regression coefficient covariance (*e. g.* 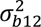) between traits is non-zero, information can be borrowed across traits, enhancing statistical power to detect gene sets associated with the traits.

Similarly, the residual covariance matrix 𝑽_𝑬_ is defined as:

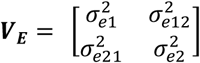

This matrix accounts for residual variance and covariance not explained by gene set effects, encompassing both trait-specific variations and measurement errors.

### Implementation of BLR model analysis

The BLR model parameter estimates (*e.g.*, 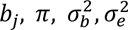 for the single trait model) were obtained using Markov Chain Monte Carlo (MCMC) Gibbs sampling procedures [36]. For analyses involving both single-trait and multi-trait scenarios, a total of 3000 iterations were employed, with the initial 500 iterations designated as burn-in to ensure adequate model convergence. Multiple runs were conducted to confirm convergence.

### Gene-level statistics

The gene-level statistics were computed as the sum of the squared SNP-level 𝑧-values 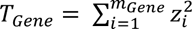 and gene-based *P*-values were calculated using the VEGAS algorithm [20]. This approach accounts for LD between SNPs by using the distribution of quadratic forms in normally distributed variables and employing saddle point approximations [37, 38], as implemented in the *vegas* function of the qgg package [36, 40]. For gene-set analysis, each gene 𝑔 has its *P*-value 𝑝_𝑔_converted to a Z-value 𝑧_𝑔_ = Φ^−1^(1 − 𝑝_𝑔_), where Φ^−1^ is the probit function. This results in a roughly normally distributed variable 𝑍, indicating the strength of the association between each gene and the trait, with higher values representing stronger associations. For the gene-level association statistics, ancestry-matched LD information for each gene region was obtained from the 1000 Genomes Project reference panel [30].

### Estimation of genetic parameters

We estimated SNP-based heritability and genetic correlations using linkage disequilibrium score regression (LDSC) [39], as implemented in the R package qgg [36, 40]. This method leverages summary statistics from GWAS to quantify the proportion of phenotypic variance attributable to common genetic variants (SNP heritability) and to assess the shared genetic architecture between traits (genetic correlation), while accounting for linkage disequilibrium and potential confounding biases such as population stratification. LD scores, as well as heritability and genetic correlation estimates, were obtained using functions provided in qgg [36, 40]. Traits included in the analysis were type 2 diabetes (T2D) [22], coronary artery disease (CAD) [23], chronic kidney disease (CKD) [24], hypertension (HTN) [25], body mass index (BMI) [26], waist-hip ratio (WHR) [26], glycated haemoglobin (HbA1c) [24], height [27], systolic blood pressure (SBP) [28], and triglycerides (TG) [29].

### Measuring the degree of enrichment

Gene sets with PIP ≥ 0.5 were considered associated. We have previously shown that our BLR procedure provides well-calibrated PIP values, accurately reflecting the probability of each gene set association with the disease [15]. Additionally, we employed another association metric from the BLR model: the posterior mean of regression effects. Gene sets with PIP>0.05 and negative regression estimates (𝒃^) were included in the analyses but not interpreted, as they may reflect enrichment of non-associated genes.

### Enrichment analysis using hypergeometric test

To support the top-ranking ATC groups and drugs identified by the BLR method by external evidence, an enrichment analysis of disease terms, using a hypergeometric test, was performed [41]. Each ATC group/drug was tested for enrichment of disease-gene associations from the DISEASES database [42, 43], with scores assessed based on curated knowledge databases, experimental data (primarily GWAS Catalogue [44]), and automated text mining of biomedical literature. Enrichment analyses were performed both collectively and separately for each information channel, including knowledge base, text mining, and experimental data. The disease term used was “Type 2 Diabetes mellitus”.

## Results

### Genetic correlations between T2D and related traits

To motivate the inclusion of multiple traits in the gene set modelling, we first examined the pairwise genetic correlations between T2D and a panel of metabolically and cardiovascular relevant phenotypes (Figure 2). As expected, T2D showed strong positive genetic correlations with BMI, HbA1c, TG, CAD, HTN, and SBP. Notably, BMI exhibited the strongest correlation with T2D, followed by HbA1c and HTN, suggesting a shared polygenic architecture between T2D and these traits. Weaker or near-zero correlations were observed between T2D and WHR or CKD, indicating more modest genetic overlap with these outcomes.

**Figure 2.**
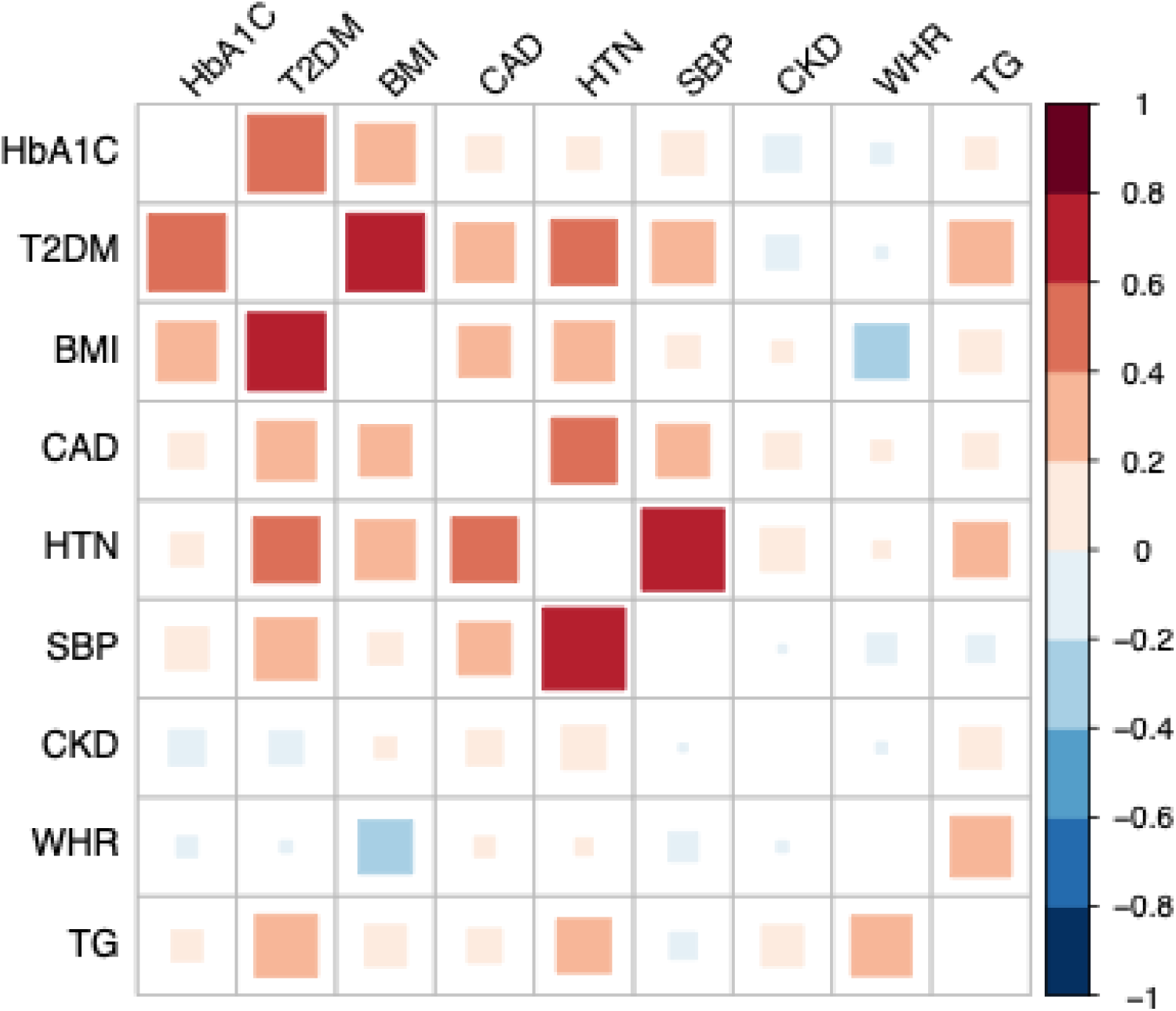
Genetic correlations between type 2 diabetes (T2D) and related cardiometabolic traits. Pairwise genetic correlations (𝑟_𝑔^_) were estimated using LD Score Regression for T2D and eight related traits: glycated haemoglobin (HbA1c), body mass index (BMI), coronary artery disease (CAD), hypertension (HTN), systolic blood pressure (SBP), chronic kidney disease (CKD), waist-hip ratio (WHR), and triglycerides (TG). The colour scale indicates the direction and magnitude of genetic correlation (red = positive; blue = negative), and square size reflects the absolute value of the correlation coefficient. Results highlight strong genetic overlap between T2D and several metabolic traits, particularly BMI, HbA1c, and HTN.

### Drug groups associated with T2D and related traits

The probability that a given drug group is relevant to the model for a specific trait, here quantified using PIP, was estimated for each drug group (ATC 4th level) with respect to T2D and related traits (Figure 3). Notably, long-acting insulins and analogues (A10AE) and other blood glucose-lowering drugs (A10BX), displayed high PIP values for T2D, underscoring the model’s ability to identify existing T2D treatments. Insulins were also strongly associated with HTN, WHR, TG, and SBP, and moderately with HbA1c. Other blood glucose-lowering drugs were linked to HbA1c, BMI, and WHR.

**Figure 3.**
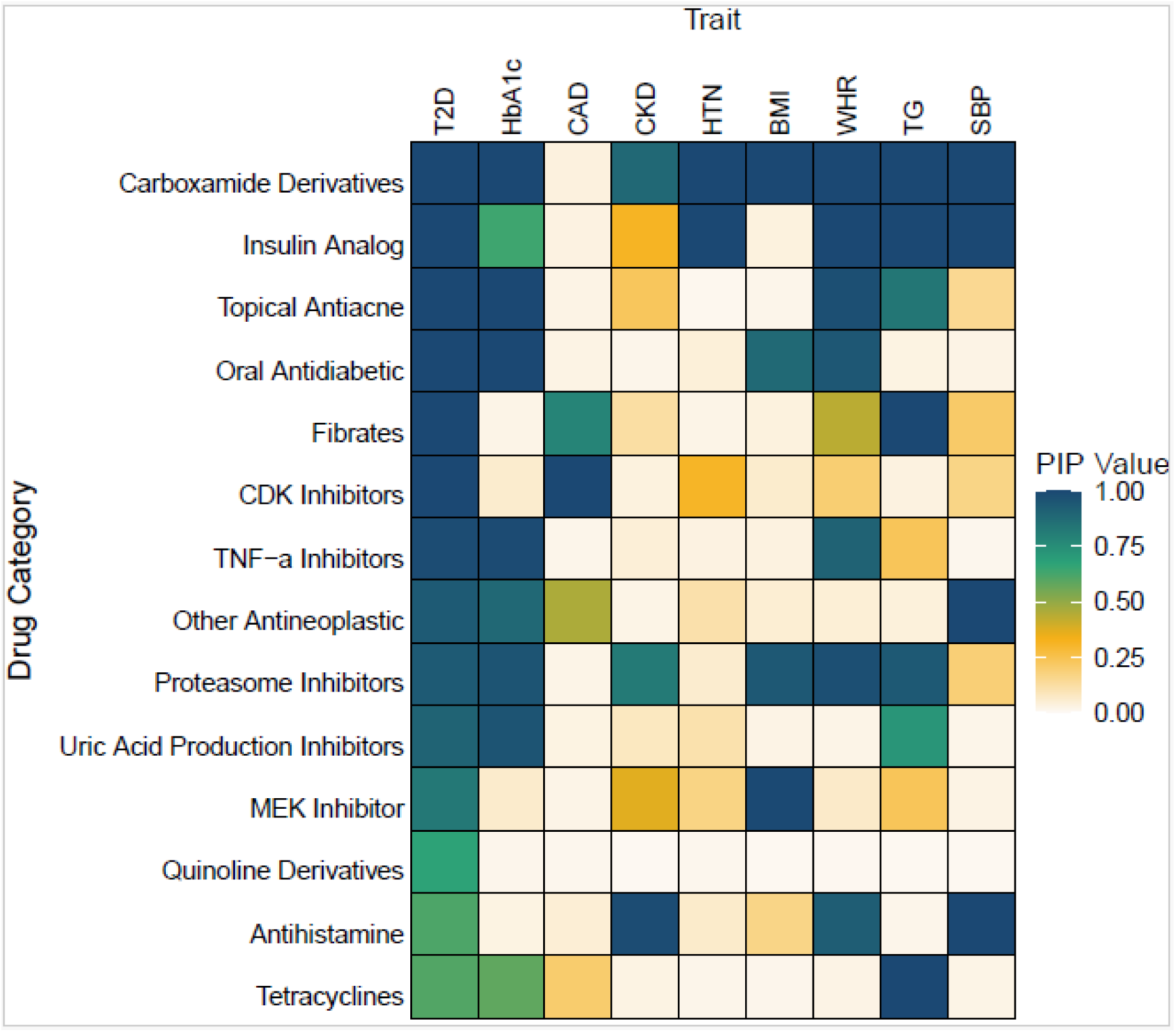
Comparative heatmap analysis of ATC 4^th^ level groups associations with type 2 diabetes and correlated traits using the multi-trait Bayesian Linear Regression (BLR) model. The heatmap visualizes the associations between ATC 4^th^ level groups and type 2 diabetes (T2D) along with correlated traits. Columns represent traits analysed through genome-wide association studies (GWAS), while rows correspond to ATC groups. The colour scale indicates the Posterior Inclusion Probability (PIP), ranging from 0.00 to 1.00. Higher PIP values denote stronger associations, suggesting a higher likelihood that the drug is relevant to the trait. Data with positive beta values and a PIP larger than 0.5 for T2D is included. Haemoglobin A1c (Hb1Ac), coronary artery disease (CAD), chronic kidney disease (CKD), hypertension (HTN), body mass index (BMI), waist-hip ratio (WHR), triglyceride (TG), systolic blood pressure (SBP).

Other drug groups associated with T2D included carboxamide derivates (N03AF), topical anti-acne preparations (D10AX), fibrates (C10AB), uric acid production inhibitors (M04AA), quinoline derivatives (P02BA), substituted alkylamines (R06AB), and tetracycline (D06AA). In addition, several antineoplastic and immunomodulating agents had high PIP values for T2D, including cyclin-dependent kinase (CDK) inhibitors (L01EF), tumour necrosis factor alpha (TNF-α) inhibitors (L04AB), proteasome inhibitors (L01XG), mitogen-activated protein kinase (MEK) inhibitors (L01EE), and other antineoplastic agents (L01XX) (Figure 3).

Multiple drug groups exhibited associations across various traits. Carboxamide derivatives, showed associations with nearly all related traits, including HbA1c, CKD, HTN, BMI, WHR, TG, and SBP. Fibrates were strongly associated with TG and coronary artery disease CAD. Uric acid production inhibitors were linked to HbA1c and TG, while substituted alkylamines were associated with CKD, WHR, and SBP. The various antineoplastic and immunomodulating agents were associated with HbA1c, CAD, CKD, BMI, WHR, TG, and SBP.

An enrichment analysis, integrating data from text mining and the GWAS catalogue, showed significant enrichment of diabetes-related genes within 9 of the 14 top ranking drug groups (Table 1). None of the drug groups were found in the knowledge base. Notably, the group of insulins were not significantly enriched with diabetes-related genes (Table 1).

**Table 1.** *P* values from enrichment analyses for the disease term “Type 2 Diabetes mellitus” based on text mining and GWAS catalog data.

| **ATC group** | **Text mining** | **GWAS catalogue** | **Together** |
| --- | --- | --- | --- |
| Carboxamide derivates (N03AF) | *P* <0.0001 | *P* =0.6 | *P* <0.0001 |
| Insulin analogues (A10AE) | *P* =0.15 | *P* =0.0001 | *P* =0.1 |
| Topical antiacne (D10AX) | *P* =0.0001 | *P* =0.5 | *P* <0.0001 |
| Blood glucose lowering drugs (A10BX) | *P* <0.0001 | *P* <0.0001 | *P* <0.0001 |
| Fibrates (C10AB) | *P* <0.0001 | *P* <0.0001 | *P* <0.0001 |
| CDK inhibitors (L01EF) | *P* <0.0001 | *P* <0.0001 | *P* <0.0001 |
| TNF-α inhibitors (L04AB) | *P* =0.03 | *P* =0.4 | *P* =0.03 |
| Other antineoplastic agents (L01XX) | *P* <0.0001 | *P* <0.0001 | *P* <0.0001 |
| Proteasome inhibitors (L01XG) | *P* <0.0001 | *P* =0.05 | *P* <0.0001 |
| Uric acid production inhibitors (M04AA) | *P* <0.0001 | *P* =0.3 | *P* <0.0001 |
| MEK inhibitors (L01EE) | *P* <0.0001 | *P* <0.0001 | *P* <0.0001 |
| Quinoline derivatives (P02BA) | *P* =0.5 | *P* =1 | *P* =0.5 |
| Antihistamines (R06AB) | *P* =0.3 | *P* =0.1 | *P* =0.3 |
| Tetracyclines (D06AA) | *P* =0.3 | *P* =1 | *P* =0.3 |

### T2D-associated genes linked to drugs within the highest-ranking drug groups

Insulins were linked to genes encoding insulin and the insulin receptor (Figure 4). Other blood glucose-lowering drugs were associated with multiple T2D-relevant genes, including *KCNQ1*, *KCNJ11*, *GCK*, *ABCC8*, and *GLP1R* - all involved in pancreatic beta-cell function, insulin secretion, or glucose sensing.

**Figure 4.**
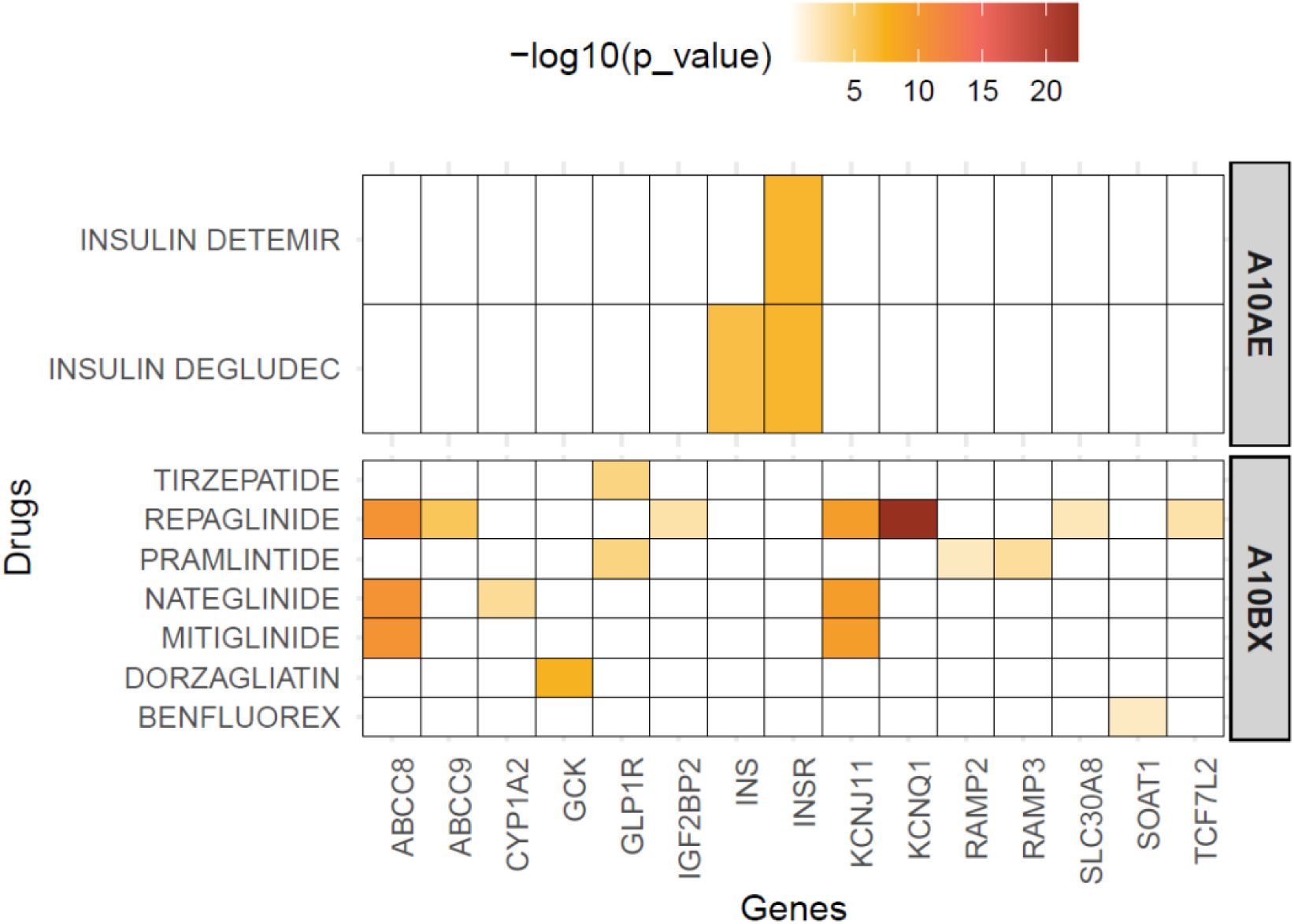
Distribution and significance of T2D-associated genes within drugs belonging to long-acting insulins and analogues (A10AE) and other blood glucose-lowering drugs (A10BX). The heatmap shows T2D-associated genes identified through GWAS using the multi-trait BLR model. Columns represent genes, and rows correspond to drugs. The colour scale indicates the negative logarithm of the *P* value (-log(*P* value)) calculated using VEGAS, with higher values reflecting stronger statistical significance and a greater likelihood of gene association with T2D. Long-acting insulins and analogues (A10AE), other blood glucose-lowering drugs (A10BX).

Also, non-glucose-lowering drugs showed associations with T2D-related genes, suggesting links to diabetes-relevant pathways (Figure 5-7). Fibrates were linked to genes involved in lipid and glucose metabolism, including *LPL, GCKR*, and *PPARG*. Carbamazepine was associated with *TNF*, *LTA*, *PSORS1C*, and *HLA−B*, *HLA−DQB1*, and *HLA−DRB1,* which also showed association to allopurinol. Carboxamide derivates showed links to genes encoding sodium voltage-gated channel proteins (*SCN11A*, *SCN1A*, *SCN2A*, *SCN7A*, and *SCN9A*) (Figure 5). Kinase inhibitors were associated with kinase-related genes, proteasome inhibitors with proteasome-related genes, and TNF-α inhibitors with *TNF* (Figure 6). The group of other antineoplastic agents showed widespread associations, including *KCNQ1*, *TNF*, and *PPARG* (Figure 7).

**Figure 5.**
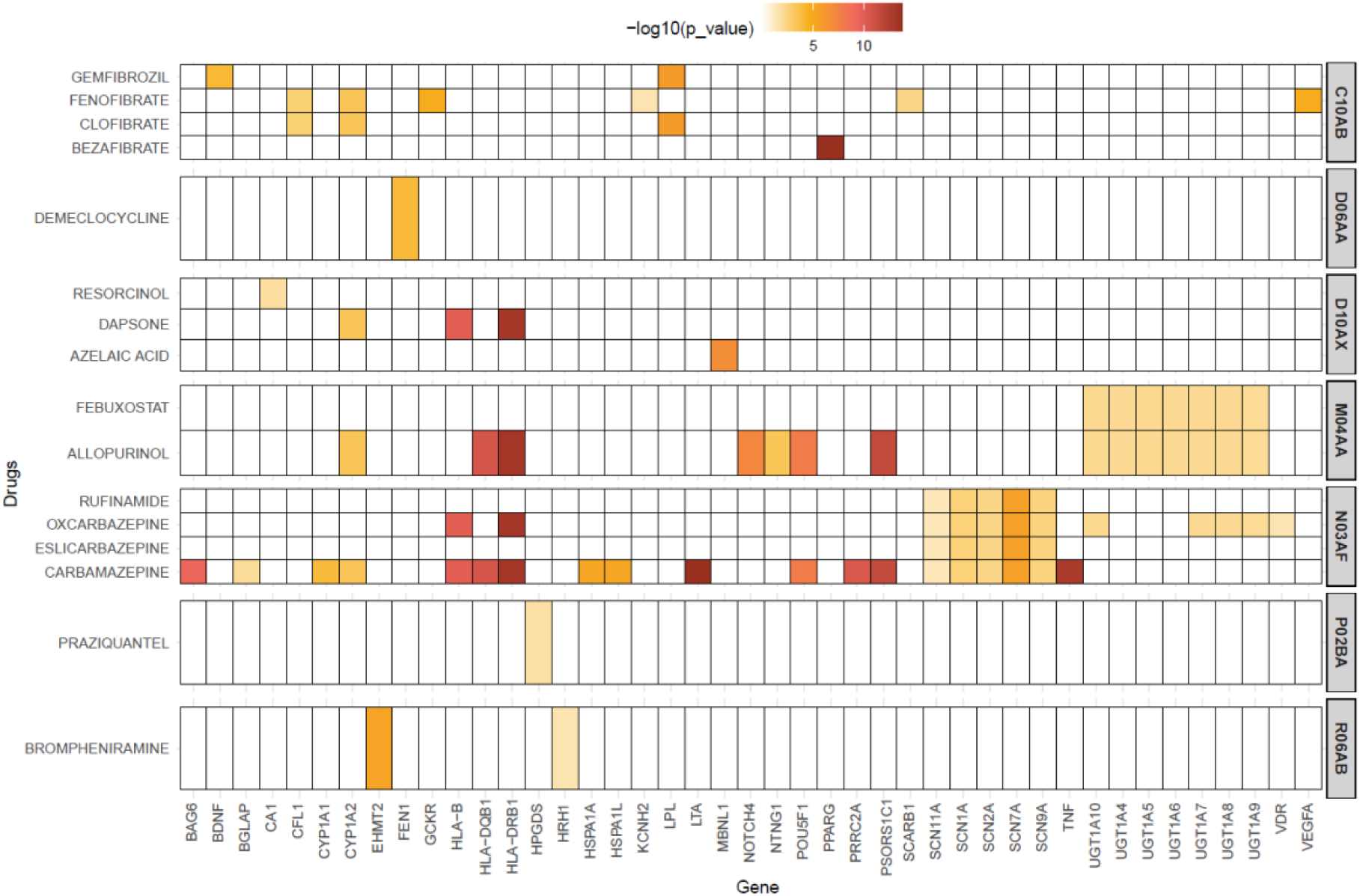
Distribution and significance of T2D-associated genes within drugs belonging to fibrates (C10AB), tetracycline (D06AA), topical anti-acne preparations (D10AX), uric acid production inhibitors (M04AA), carboxamide derivates (N03AF), quinoline derivatives (P02BA), and substituted alkylamines (R06AB). The heatmap shows T2D-associated genes identified through GWAS using the multi-trait BLR model. Columns represent genes, and rows correspond to drugs. The colour scale indicates the negative logarithm of the *P* value (-log(*P* value)) calculated using VEGAS, with higher values reflecting stronger statistical significance and a greater likelihood of gene association with T2D. Fibrates (C10AB), tetracycline (D06AA), topical anti-acne preparations (D10AX), uric acid production inhibitors (M04AA), carboxamide derivates (N03AF), quinoline derivatives (P02BA), and substituted alkylamines (R06AB).

**Figure 6.**
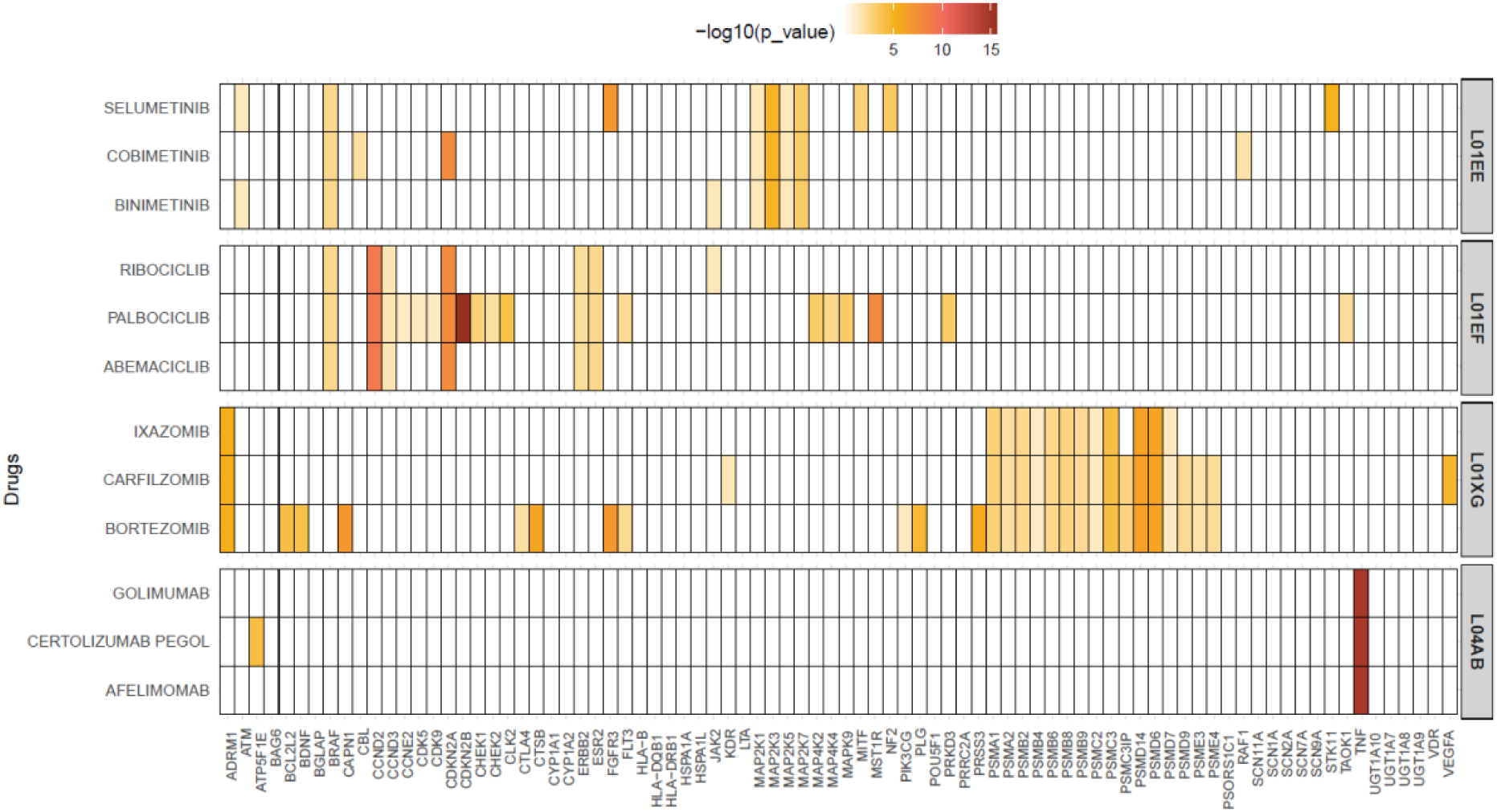
Distribution and significance of T2D-associated genes within drugs belonging to MEK inhibitors (L01EE), CDK inhibitors (L01EF), proteasome inhibitors (L01XG), and TNF-α inhibitors (L04AB). The heatmap shows T2D-associated genes identified through GWAS using the multi-trait BLR model. Columns represent genes, and rows correspond to drugs. The colour scale indicates the negative logarithm of the *P* value (-log(*P* value)) calculated using VEGAS, with higher values reflecting stronger statistical significance and a greater likelihood of gene association with T2D. Cyclin-dependent kinase (CDK) inhibitors (L01EF), tumour necrosis factor alpha (TNF-α) inhibitors (L04AB), proteasome inhibitors (L01XG), mitogen-activated protein kinase (MEK) inhibitors (L01EE).

**Figure 7.**
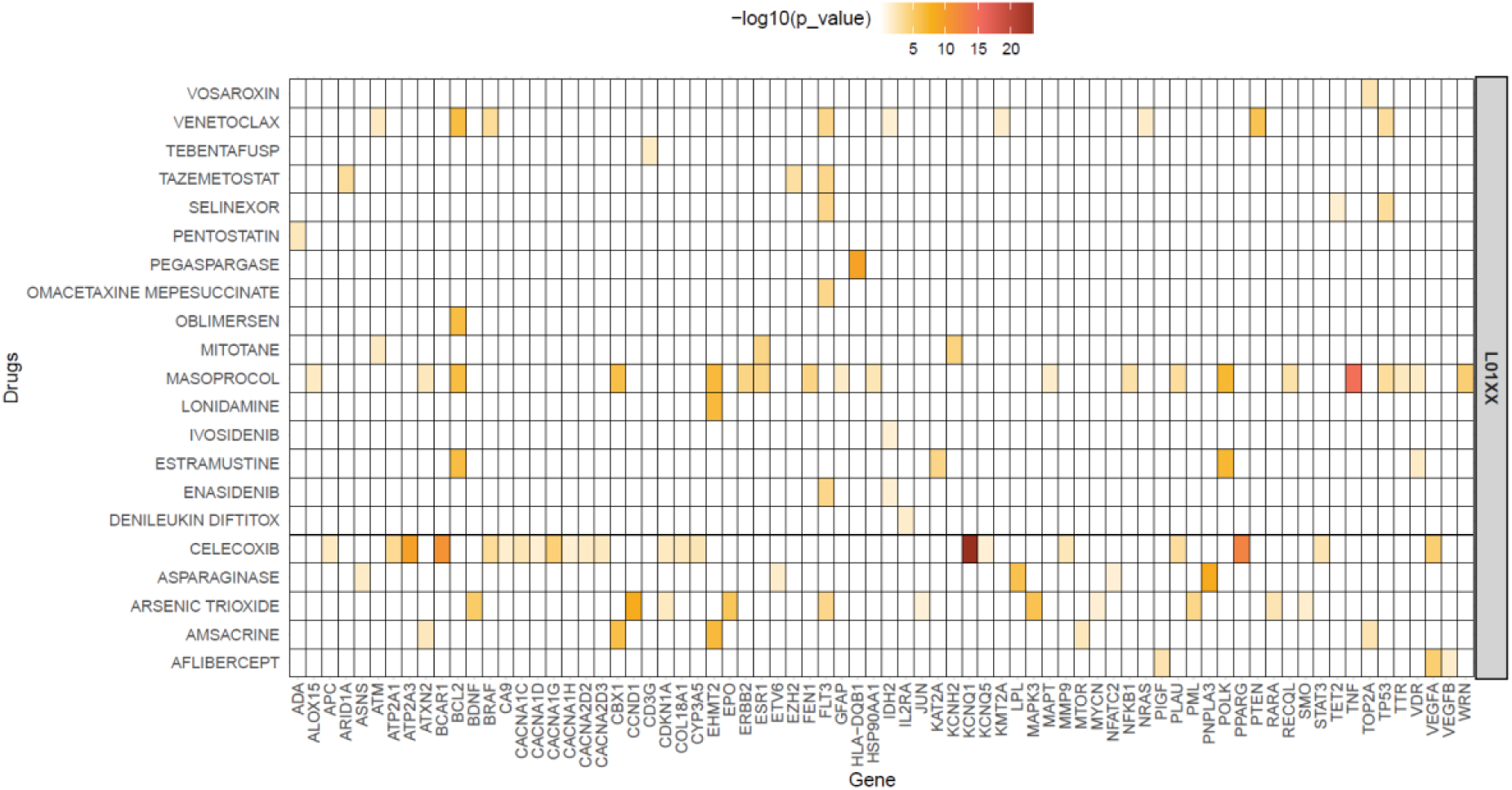
Distribution and significance of T2D-associated genes within drugs belonging to other antineoplastic agents (L01XX). The heatmap shows T2D-associated genes identified through GWAS using the multi-trait BLR model. Columns represent genes, and rows correspond to drugs. The colour scale indicates the negative logarithm of the *P* value (-log(*P* value)) calculated using VEGAS, with higher values reflecting stronger statistical significance and a greater likelihood of gene association with T2D. Other antineoplastic agents (L01XX).

## Discussion

Here we present a novel genetically informed drug-repurposing strategy for T2D, utilizing the BLR multi-trait model to identify drugs displaying genetic association with T2D genetic architecture. This approach integrates GWAS summary statistics from a comprehensive T2D study with drug-gene interaction data sourced from DGIdb. The model successfully identified established as well as new associations, which supports the model’s effectiveness and indicates the potential of non-traditional drugs in managing T2D and its associated conditions.

As expected, blood glucose-lowering drugs showed a high enrichment of genes linked to T2D and HbA1c, reflecting the close link between these drugs and the genetic factors of the disease. This supports the model’s ability to identify potential drugs relevant for T2D management.

Several other drug groups were linked to T2D-associated genes as well, including fibrates, carboxamide derivatives, uric acid production inhibitors and several immunomodulatory agents.

Fibrates, used to treat hypertriglyceridemia, was associated not only with T2D but also with triglyceride levels and coronary artery disease. Bezafibrate showed a distinct association with T2D, likely due to its unique action profile. Unlike other fibrates that target only PPAR-α, a key regulator of lipid metabolism, bezafibrate also activates PPAR-γ and -δ [45]. PPAR-γ, genetically linked to T2D, influences adipogenesis, lipid metabolism, glucose homeostasis, and inflammation. While selective PPAR-γ agonists (e.g., glitazones) improve insulin sensitivity, their use is limited by adverse effects such as congestive heart failure and osteoporosis [46, 47] - side effects not associated with bezafibrate. Clinical studies have shown that bezafibrate improves insulin sensitivity and lowers blood glucose, particularly in individuals with elevated triglycerides. Moreover, bezafibrate may reduce the incidence and delay the onset of T2D in high-risk populations [45, 48–50]. The combined PPAR-α/-γ action may simultaneously target insulin resistance and atherogenic dyslipidaemia [2,6], while PPAR-δ is suggested to support weight regulation [45, 51]. Bezafibrate may therefore offer added glycemic benefits for patients with pre-diabetes and dyslipidemia, beyond those seen with conventional lipid-lowering therapies.

Carboxamide derivatives, particularly carbamazepine, were associated with several genes strongly linked to T2D. Isolated case reports have described hyperglycaemia or new-onset diabetes following carbamazepine overdose [52]; however, data from a large cohort study do not support a diabetogenic effect at therapeutic doses [53]. Moreover, preclinical studies in Non-Obese Diabetic (NOD) mice have shown that carbamazepine preserves pancreatic β-cell function and improves glycaemic regulation, potentially delaying or reducing the onset of type 1 diabetes [54].

Allopurinol was also enriched for T2D-associated genes. Growing evidence suggests that uric acid contributes to oxidative damage, and hyperuricemia has been associated with increased risk of HTN, CKD, cardiovascular disease, metabolic syndrome, and type 2 diabetes. [55]. Small clinical studies have reported improved insulin sensitivity, as measured by HOMA-IR, and reduced levels of high-sensitivity C-reactive protein in hyperuricemic individuals, both with and without T2D, following allopurinol treatment [56, 57]. In addition, allopurinol may confer cardiovascular benefits in patients with T2D, potentially through reductions in inflammation, oxidative stress, and improvements in glycemic and lipid profiles [58]. However, evidence for glycemic benefit remains inconclusive [55], and large-scale clinical trials are needed to clarify allopurinol’s role in the prevention and treatment of T2D.

The observed association between T2D-associated genes and immunomodulatory and antineoplastic agents underscores the role of inflammation in the pathophysiology of type 2 diabetes [59]. Anti-inflammatory therapies, such as TNF-α inhibitors, have accordingly been proposed as potential strategies to target insulin resistance and T2D [60–63], although most supporting evidence to date stems from preclinical studies.

Genetically informed drug repurposing is increasingly applied, particularly for complex diseases such as T2D. Prior approaches have leveraged Mendelian randomization and s-PrediXcan-based gene expression estimates from GTEx and GWAS data, combined with drug–gene mappings from resources such as DGIdb, to identify candidate drug classes, including antihypertensives and lipid-lowering agents [64, 65]. In contrast, our study employed a multi-trait BLR model that integrates a broader set of T2D-relevant traits to enhance statistical power and gene set prioritization [15, 66]. We additionally applied curated gene sets at the 4th-level ATC classification, enabling more granular analysis of drug classes and potential mechanisms. To reduce false positives, we incorporated rigorous LD correction using the VEGAS algorithm. Together, this framework supports scalable and flexible evaluation of overlapping gene sets across multiple traits, offering a complementary strategy for genetically informed repurposing.

A key limitation of this study is the reliance on DGIdb. Although DGIdb is a widely used and well-curated resource, it aggregates interactions from heterogeneous sources with varying levels of evidence. Consequently, some gene–drug associations may be incomplete, inaccurate, or context-dependent. This introduces uncertainty in the downstream analyses, as erroneous or non-specific drug–gene links can lead to spurious signals or obscure true repurposing opportunities.

## Conclusion

Drug repurposing offers a faster and more cost-effective alternative to de novo drug development, as safety and pharmacokinetic profiles are already established [10]. This study demonstrates the potential of a multi-trait BLR approach for identifying genetically supported targets in T2D. By integrating correlated traits, curated gene sets, and rigorous LD correction, the model enables scalable and robust prioritization of repurposing candidates. As genetic heterogeneity in T2D becomes increasingly evident, such approaches may also support precision medicine by aligning drug selection with individual genetic profiles. We identified fibrates, carboxamide derivatives, uric acid inhibitors, and immunomodulatory agents as potential therapeutic targets for T2D. These findings warrant further investigation, including evaluation of selected candidates in future randomized controlled trials.

## Funding

This project was supported by the Novo Nordisk Foundation through the Open Discovery Innovation Network (ODIN) drug discovery platform under grant number NNF20SA0061466. The funding initiative aims to foster collaboration between academia and industry, promoting innovative research with long-term impact.

## Data availability

The datasets analysed in this study are publicly available from the following sources:

- GWAS summary statistics were obtained from the Type 2 Diabetes Knowledge Portal (https://t2d.hugeamp.org/datasets.html), including:

- CARDIoGRAMplusC4D-UK Biobank CAD 2018 GWAS Meta-analysis
- CKDGen 2019 GWAS
- Heart Rate and Hypertension GWAS
- GIANT-UK Biobank GWAS Meta-analysis
- UK Biobank 2021 HbA1c GWAS (European ancestry)
- UKB and ICBP Blood Pressure GWAS
- ERA Additive Model Age-Related GWAS
- Disease-gene associations were sourced from DISEASES: https://diseases.jensenlab.org
- 1000 Genomes Project reference data for European, East Asian, and South Asian populations were downloaded from the Centre for Neurogenomics and Cognitive Research (CNCR) at https://cncr.nl/research/magma/ (files: g1000eur.zip, g1000eas.zip, g1000sas.zip).
- The BLR prioritization method used in this study is implemented in the open source qgg R package: https://psoerensen.github.io/qgg.
- Example scripts and tutorials for applying the method are available at: https://psoerensen.github.io/gact.

## Competing Interests

The authors declare that they have no conflicts of interest related to this work.

## Notes

### Competing Interest Statement

The authors have declared no competing interest.

### Author Declarations

GWAS summary statistics were obtained from the Type 2 Diabetes Knowledge Portal (https://t2d.hugeamp.org/datasets.html), including: - CARDIoGRAMplusC4D-UK Biobank CAD 2018 GWAS Meta-analysis - CKDGen 2019 GWAS - Heart Rate and Hypertension GWAS - GIANT-UK Biobank GWAS Meta-analysis - UK Biobank 2021 HbA1c GWAS (European ancestry) - UKB and ICBP Blood Pressure GWAS - ERA Additive Model Age-Related GWAS - Disease-gene associations were sourced from DISEASES: https://diseases.jensenlab.org 1000 Genomes Project reference data for European, East Asian, and South Asian populations were downloaded from the Centre for Neurogenomics and Cognitive Research (CNCR) at https://cncr.nl/research/magma/ (files: g1000eur.zip, g1000eas.zip, g1000sas.zip). The BLR prioritization method used in this study is implemented in the open source qgg R package: https://psoerensen.github.io/qgg. Example scripts and tutorials for applying the method are available at: https://psoerensen.github.io/gact.

